# Sodium-glucose Cotransporter-2 Inhibitors Stabilize Coronary Plaques in Acute Coronary Syndrome with Diabetes Mellitus

**DOI:** 10.1101/2023.07.31.23293462

**Authors:** Atsumasa Kurozumi, Koki Shishido, Takayoshi Yamashita, Daisuke Sato, Syuhei Uchida, Eiji Koyama, Yusuke Tamaki, Takahiro Hayashi, Hirokazu Miyashita, Hiroaki Yokoyama, Tomoki Ochiai, Masashi Yamaguchi, Noriaki Moriyama, Kazuki Tobita, Takashi Matsumoto, Shingo Mizuno, Futoshi Yamanaka, Yutaka Tanaka, Masato Murakami, Saeko Takahashi, Shigeru Saito

## Abstract

**Background:** Sodium-glucose cotransporter-2 inhibitors (SGLT2i) are widely used in cardiology and are effective in treating acute coronary syndrome (ACS). Their effects on unstable plaque in ACS patients remains unclear. This study aimed to examine the effectiveness of SGLT2i in coronary plaque phenotypes based on optical coherence tomography (OCT) images and the prognosis of ACS with type 2 diabetes mellitus (T2DM).

**Methods:** This retrospective study included 109 patients in the total cohort and 29 patients in the OCT cohort. Based on SGLT2i administration after ACS, the total cohort was categorized into non-SGLT2i (n = 69) and SGLT2i (n = 40) groups. The OCT cohort had 15 and 14 patients in non-SGLT2i and SGLT2i groups, respectively. OCT images of unstable plaque were analyzed in non-stented lesions during ACS catheterization and at 6-month follow-ups. The total cohort was assessed after 1 year for major adverse cardiovascular events (MACE), including all-cause mortality, revascularization, cerebrovascular disease, and heart failure hospitalization.

**Results:** SGLT2i improved unstable lesions with a significantly thicker fibrous cap (48 ± 15 μm vs. 26 ± 24 μm, p = 0.005), reduced lipid arc (-29 ± 12° vs. -18 ± 14°, p = 0.028), and higher % decrease in total lipid arc (-35 ± 13% vs. -19 ± 18%, p = 0.01) as well as a lower MACE incidence (Log rank p = 0.023, HR 4.72 [1.08, 20.63]) and revascularization rate (adjusted HR 6.77 [1.08, 42.52]) compared to the non-SGLT2i group.

**Conclusions:** SGLT2i stabilizes atherosclerosis and improves ACS prognosis in patients with T2DM.

**Clinical Perspective:** *What is new?:*

- We analyzed serial OCT images both baseline and follow-up in patients with ACS and T2DM undergoing PCI, and evaluated how effect SGLT2i had on unstable plaques.
- ACS patients with SGLT2i administration showed coronary atherosclerotic lesions to be stabilized on follow-up.

*What are the clinical implications?:*

- SGLT2i can stabilize unstable plaques by controlling the inflammatory cascade on patients with T2DM.
- SGLT2i administration will be one option to improve their prognosis in patients with ACS and T2DM undergoing PCI.

## INTRODUCTION

Patients with acute coronary syndrome (ACS) undergoing percutaneous coronary intervention (PCI) have a poor prognosis compared to patients with chronic coronary syndrome despite recent advances in medications, such as statins and antiplatelet agents, and device technologies, such as drug-eluting stents. In particular, patients with high coronary risk factors such as diabetes mellitus, occasionally show restenosis or de-novo lesion progression, which is difficult to prevent.^1–4^

Sodium-glucose cotransporter-2 inhibitors (SGLT2i) are drugs that inhibit sodium-glucose channels in the renal proximal tubules.^5, 6^ SGLT2i have been shown to have a primary preventive effect against cardiovascular events in patients with type 2 diabetes mellitus (T2DM).^7, 8^ Some studies have shown that SGLT2i improved a prognosis of heart failure with reduced and preserved ejection fraction.^9–11^ SGLT2i have become essential drugs for treating heart failure.^12^ The cardioprotective effect of these drugs are attributed to a variety of mechanisms, not only sodium diuresis.^13^ Recent studies have demonstrated that SGLT2i administered to patients helped to prevent myocardial infarction in patients with T2DM,^14, 15^ and to prevent subsequent cardiovascular events after myocardial infarction.^16–18^ Based on animal studies showing reduced aortic plaque lesions,^19–21^ SGLT2i are expected to be effective in treating atherosclerosis. On T2DM patients with stable angina pectoris, SGLT2i was reported to ameliorate coronary artery lesions.^22^ However, their effects on coronary artery lesions in patients with ACS have not been reported. Moreover, few studies have investigated their effects on the clinical outcomes in patients with ACS without heart failure symptoms.

This study aimed to investigate the effects of SGLT2i on the histological changes in coronary artery plaques using optical coherence tomography (OCT) and on the clinical events in patients with T2DM after ACS.

## METHODS

### Study design, setting, and population

This retrospective cohort study included 580 patients who underwent primary PCI for ACS between March 2019 and December 2021 at the Shonan Kamakura General Hospital, Japan. The study flow chart is shown in Figure 1. T2DM, defined as previously diagnosed or with HbA1c ≥ 6.5% and blood glucose ≥ 200 mg/dL on admission,^23^ coexisted in 133 patients.

**Figure 1.**
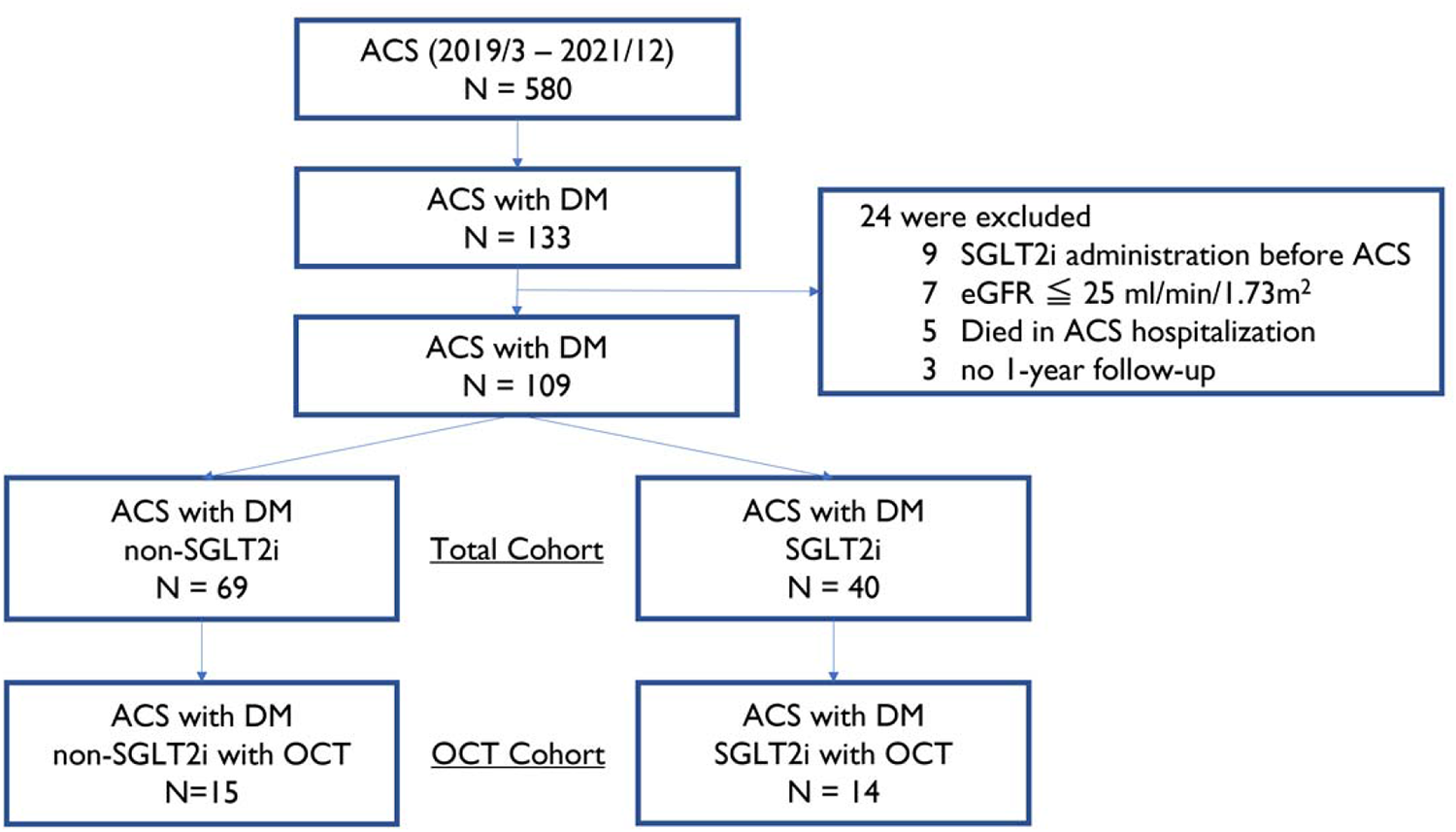
Flow chart of cohort formation. The chart shows the patient selection and exclusion criteria and the number of patients in each cohort.

We excluded patients who (a) had an estimated glomerular filtration rate (eGFR) ≤ 25 ml/min/1.73 m^2^, (b) were treated with SGLT2i before ACS hospitalization, (c) died during ACS hospitalization and (d) could not be followed up at one year after ACS. Consequently, 109 patients (69 non-SGLT2i and 40 SGLT2i) were included in the total cohort. For OCT analysis, we included 29 patients (OCT cohort: 15 non-SGLT2i and 14 SGLT2i) with OCT images of the culprit lesion at the follow-up catheterization six months after ACS catheterization (Figure 1).

Participant data were obtained from our database. The authors vouch for the completeness and accuracy of the data and all analyses. The study protocol was approved by the Mirai Medical Center Ethics Committee (TGE02092-024) and was conducted in accordance with the tenets of the Declaration of Helsinki. Informed consent was exempted using an opt-out form.

### Imaging study

We analyzed the OCT images twice for each patient. The first was at the primary PCI (baseline), and the second was at the follow-up catheterization six months later (follow-up).

We selected non-stented lesions with 1 mm slices from the distal and proximal sites of the stent edge at ACS catheterization (baseline) and at the follow-up OCT (follow-up) corresponding to each image as a reference for the side branch (Supplemental Figure 1). Slices unsuitable for analysis, such as those with insufficient blood cell removal, were excluded.

Unstable coronary lesions were defined as indistinguishable three-layer structures (intima, media, and adventitia), high signal intensity in the fibrous capsule due to macrophage accumulation, and attenuated OCT signal over the fibrous capsule. Two cardiologists calculated the fibrous cap thickness (FCT) and lipid arc on the OCT images (Ultreon™ 1.0 Software, Dragonfly™ Imaging Catheters, and the OPTIS™ Imaging Systems by Abbott Laboratories [United States]) (Figure 2). The lipid length was defined as the number of slices with unstable coronary lesions.

**Figure 2.**
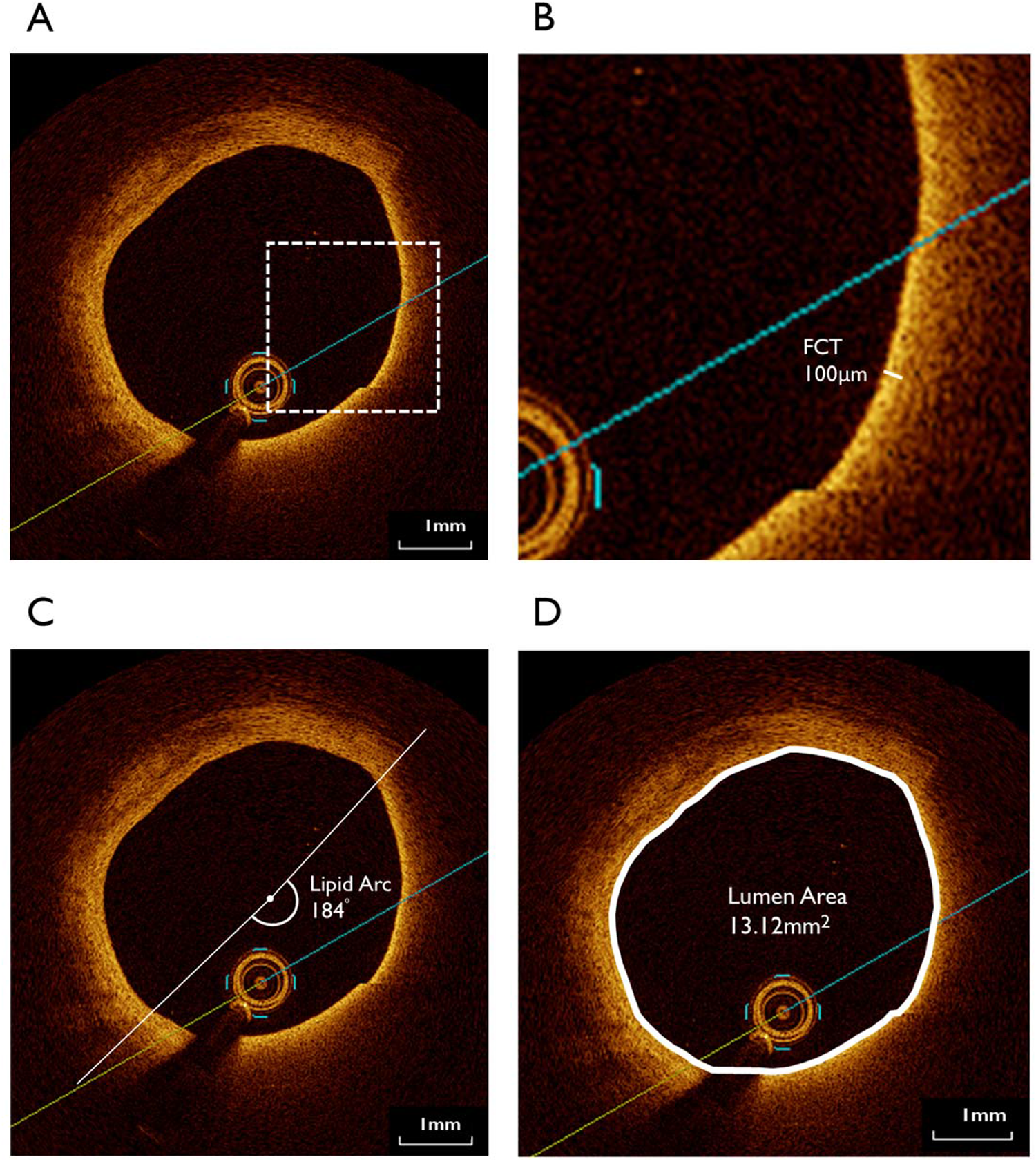
OCT measurement. OCT imaging was used to (A) confirm the presence of unstable lipid plaques, (B) measure the fibrous cap thickness, (C) measure the lipid arc, (D) measure the lumen area.

To evaluate unstable coronary lesions, we compared the rate of change in the FCT (Figure 2B), lipid arc (Figure 2C), lumen area (Figure 2D), and lipid length from baseline to follow-up between the non-SGLT2i and SGLT2i groups. FCT change was calculated as follows: minFCT_follow_ – minFCT_base_, where minFCT_base_ and minFCT_follow_ are the minimum FCT in all slices at baseline and follow-up, respectively. Similarly, lipid arc change was calculated as maxARC_follow_ – maxARC_base_, where maxARC_base_ and maxARC_follow_ are the maximum lipid arc in all slices at baseline and follow-up, respectively. Lipid arc change was also measured as % change in total lipid arc on all slices from baseline (totalARC_base_) to follow-up (totalARC_follow_): ΔtotalARC/totalARC_base_ ×100 (ΔtotalARC= totalARC_follow_ – totalARC_base_). Minimum lumen area change was calculated as follows: minLA_follow_ – minLA_base_, where minLA_base_ and minLA_follow_ are the minimum LA in all slices at baseline and follow-up, respectively. Mean lumen area change was also defined as follows: meanLA_follow_ – meanLA_base_, where meanLA_base_ and meanLA_follow_ are the mean LA in all slices at baseline and follow-up. The lipid length change was defined as follows: LL_follow_ – LL_base_, where LL_base_ and LL_follow_ are the lipid length at baseline and follow-up.

### Clinical outcome assessment

To evaluate the prognostic effect of SGLT2i, we analyzed the clinical outcomes in the non-SGLT2i and SGLT2i groups one year after ACS. Clinical outcomes included all-cause mortality, revascularization (ACS or coronary lesion progression), cerebrovascular disease, and hospitalization for heart failure, defined as major adverse cardiovascular events (MACE).

### Statistical analysis

The SGLT2i and non-SGLT2i groups were compared for categorical variables using the χ2 test and for continuous variables using paired or unpaired t-test. Univariate analysis of atherosclerosis-related variables was performed using the paired or unpaired t-test for categorical variables and the Pearson correlation test for continuous variables. The cumulative incidence of MACE was evaluated using the Kaplan–Meier curve and proportional hazards models. Baseline characteristics exhibiting P value <0.05 in the univariate analysis were included in a logistic regression analysis to determine the predictive factors of the incidence of 1-year MACE. We evaluated the incidence rates of cardiac death, non-cardiac death, revascularization, hospitalization for heart failure, and stroke using the log-rank test and unadjusted or adjusted hazard ratios using proportional hazards models. P value < 0.05 indicated statistical significance. JMP Pro® 16 (SAS Institute, Cary, North Caronia) was used for the analyses.

## RESULTS

### Baseline

Table 1 shows the baseline characteristics of patients in the total and OCT cohorts. In the total cohort, the mean age was significantly lower in the SGLT2i group (65.5 ± 13.5 years) than in the non-SGLT2i group (73.8 ± 11.8 years) (p = 0.001). HbA1c values were significantly higher in the SGLT2i than in the non-SGLT2i group at ACS hospitalization (8.7 ± 2.2 % vs. 7.2 ± 1.2 %, p < 0.001) but not at follow-up (7.2 ± 0.8 % vs. 7.0 ± 0.7 %, p = 0.278). The eGFR values were significantly higher in the SGLT2i group than in the non-SGLT2i both at baseline (67.9 ± 18.6 ml/min/1.73 m^2^ vs. 57.9 ± 17.4 ml/min/1.73 m^2^, p = 0.006) and at follow-up (63.5 ± 18.8 ml/min/1.73 m^2^ vs. 55.1 ± 13.9 ml/min/1.73 m^2^, p = 0.02).

**Table 1.**
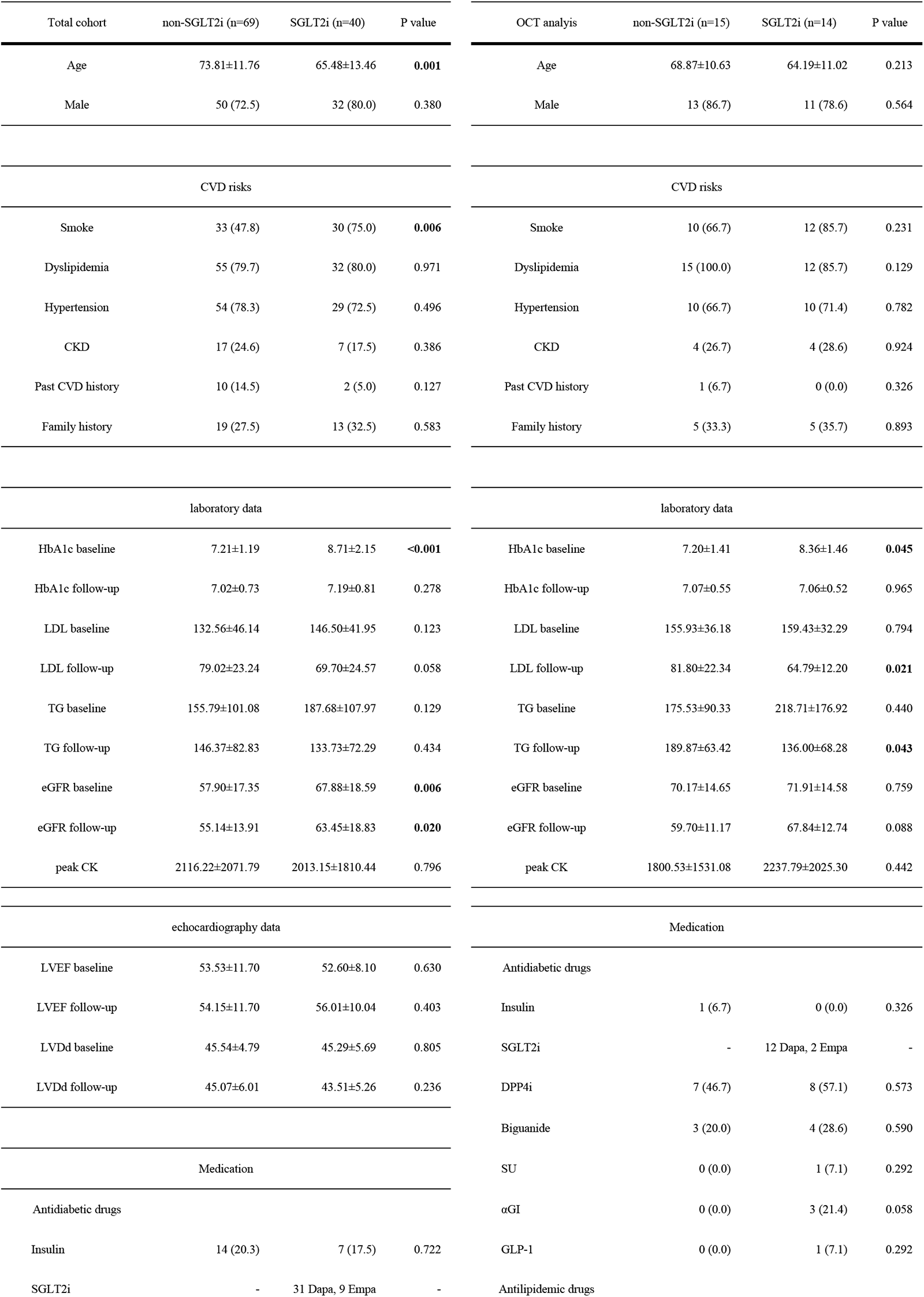
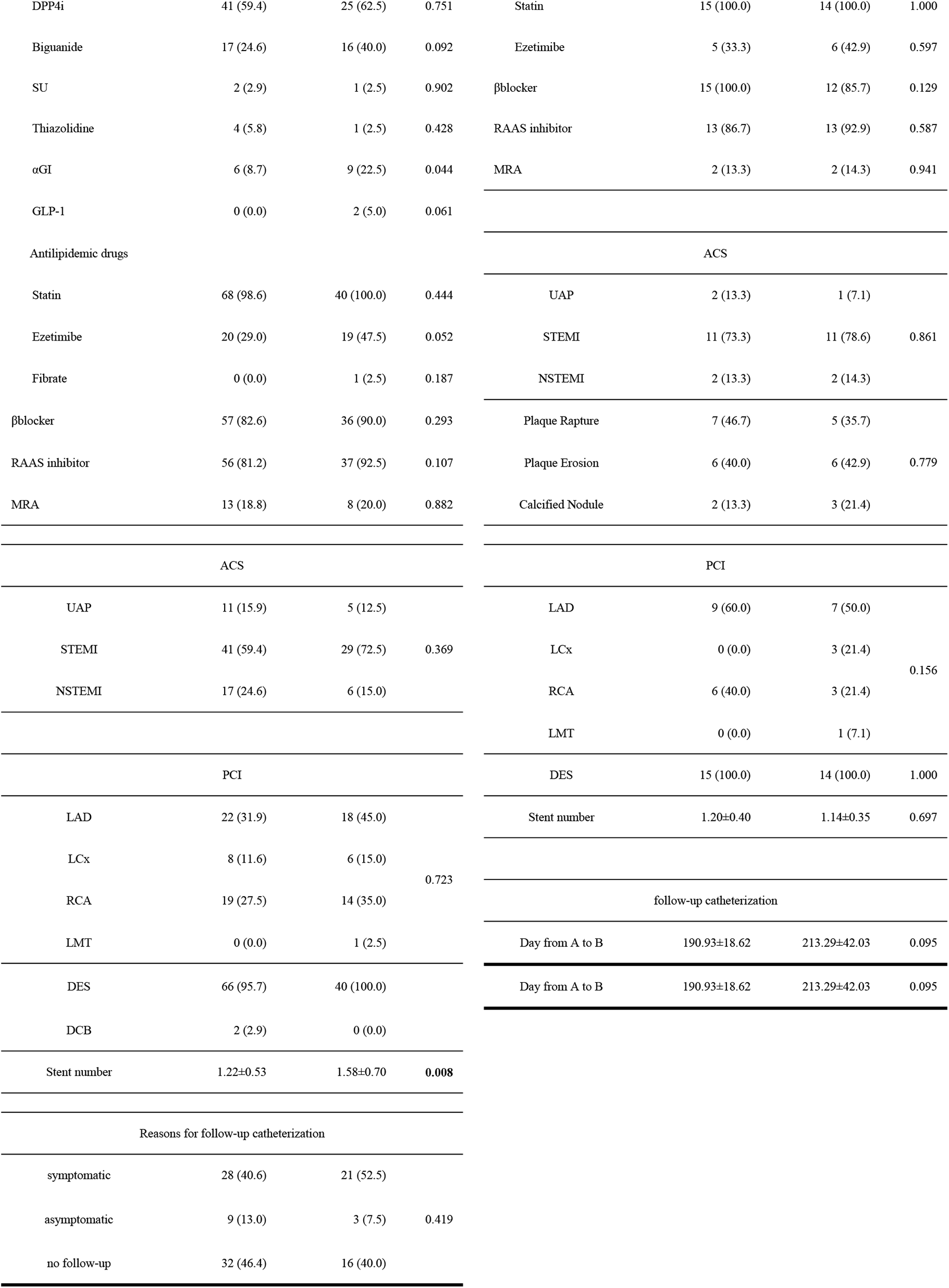
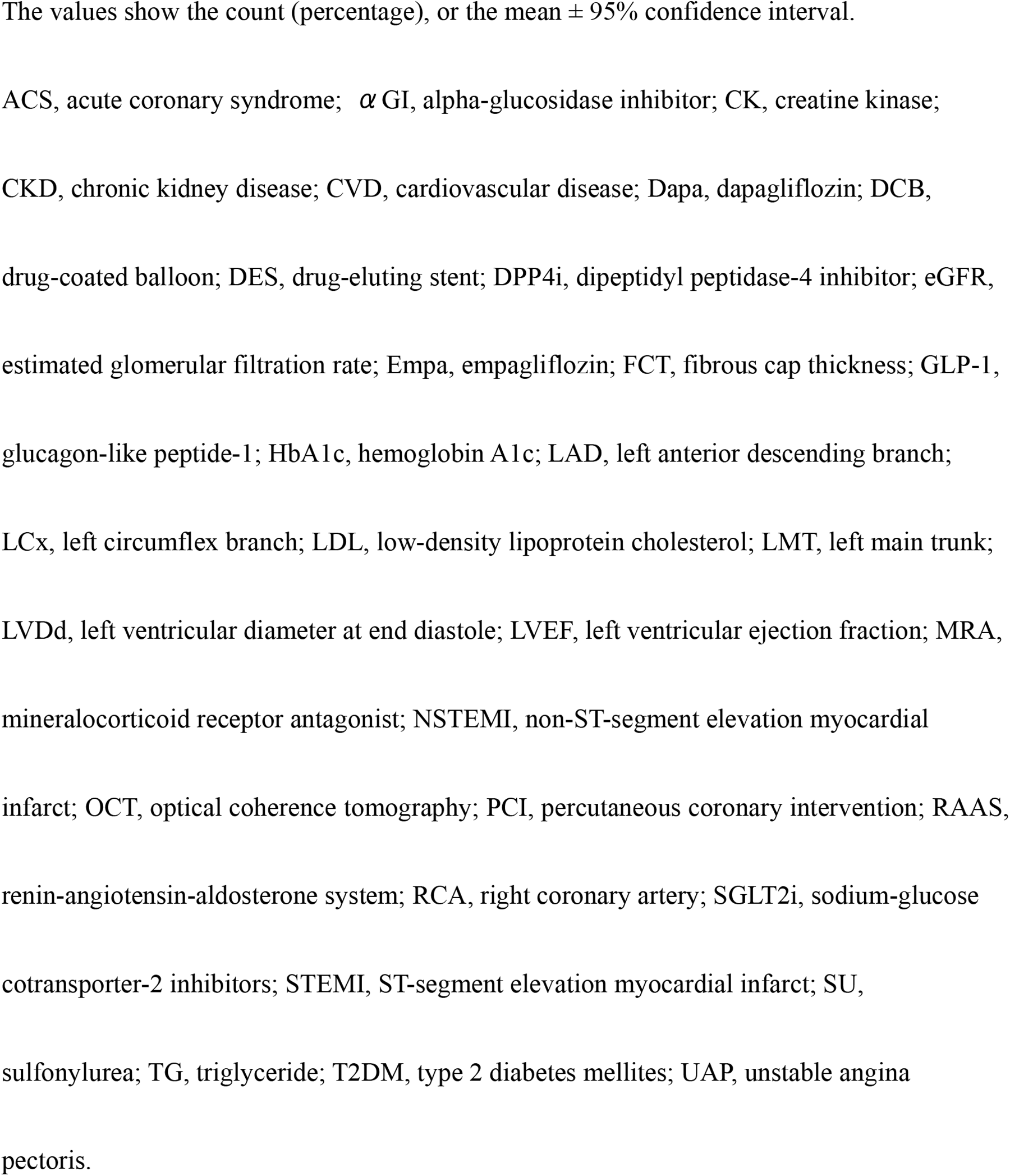
Baseline characteristics of ACS patients with T2DM.

In the OCT cohort, the SGLT2i and non-SGLT2i groups showed no significant differences in the baseline characteristics, including age, sex, risk for cardiovascular disease, and medications. The HbA1c values were significantly higher in the SGLT2i group (8.4 ± 1.5 % vs. 7.2 ± 1.4 %, p = 0.045) at ACS hospitalization but not at follow-up (7.1 ± 0.6 % vs. 7.1 ± 0.5 %, p = 0.965). The LDL cholesterol and TG at baseline showed no differences between the two groups. However, at the follow-up, the SGLT2i group had significantly lower LDL cholesterol (64.8 ± 12.2 mg/dL vs. 81.8 ± 22.3 mg/dL, p = 0.021) and TG (136.0 ± 68.3 mg/dL vs. 189.9 ± 63.4 mg/dL, p = 0.043).

### OCT analysis

To assess the effectiveness of SGLT2i for atherosclerosis, we evaluated the histological changes in the coronary artery plaque distal and proximal from the stent edge at ACS catheterization (baseline) and at follow-up by OCT (Figure 3). The period from baseline to follow-up in the OCT analysis showed no difference between the non-SGLT2i and SGLT2i groups (191 ± 19 days vs. 213 ± 42 days, p = 0.095). As shown in Table 2, the minimum FCT, maximum lipid arc, minimum/mean lumen area and lipid length at ACS catheterization were not significantly different. However, the minimum FCT at follow-up was thicker in the SGLT2i group (154 ± 44 μm vs. 122 ± 30 μm, p = 0.037). Although the maximum lipid arc was numerically larger in the SGLT2i group, the difference between the two groups was not significant at follow-up (76 ± 47 ° vs. 58 ± 36 °, p = 0.272). The minimum lumen area had no difference at follow-up while the SGLT2i group had larger lumen area (3.96 ± 2.36 mm^2^ vs. 3.52 ± 1.54 mm^2^, p=0.569). The mean lumen area was the same as well (7.20 ± 3.23 mm^2^ vs. 6.71 ± 2.37 mm^2^, p=0.567). The lipid length had no difference at follow-up between two groups (9.43 ± 6.10 mm vs. 11.20 ± 5.97 mm, p=0.452). The SGLT2i group had a significantly greater absolute change in min FCT (48 ± 15 μm vs. 26 ± 24 μm, p = 0.005), max lipid arc (-29 ± 12 ° vs. -18 ± 14 °, p = 0.028), and % change in the total lipid arc (-35 ± 13 % vs. -19 ± 18 %, p = 0.01). The two groups showed no significant differences on the absolute change in minimum lumen area (0.29 ± 1.41 mm^2^ vs. -0.03 ± 1.56 mm^2^, p=0.582), mean lumen area (0.39 ± 1.14 mm^2^ vs. -0.09 ± 0.93 mm^2^, p=0.242), and lipid length (-1.14 ± 1.85 mm vs. -1.07 ± 2.38 mm, p=0.927).

**Figure 3.**
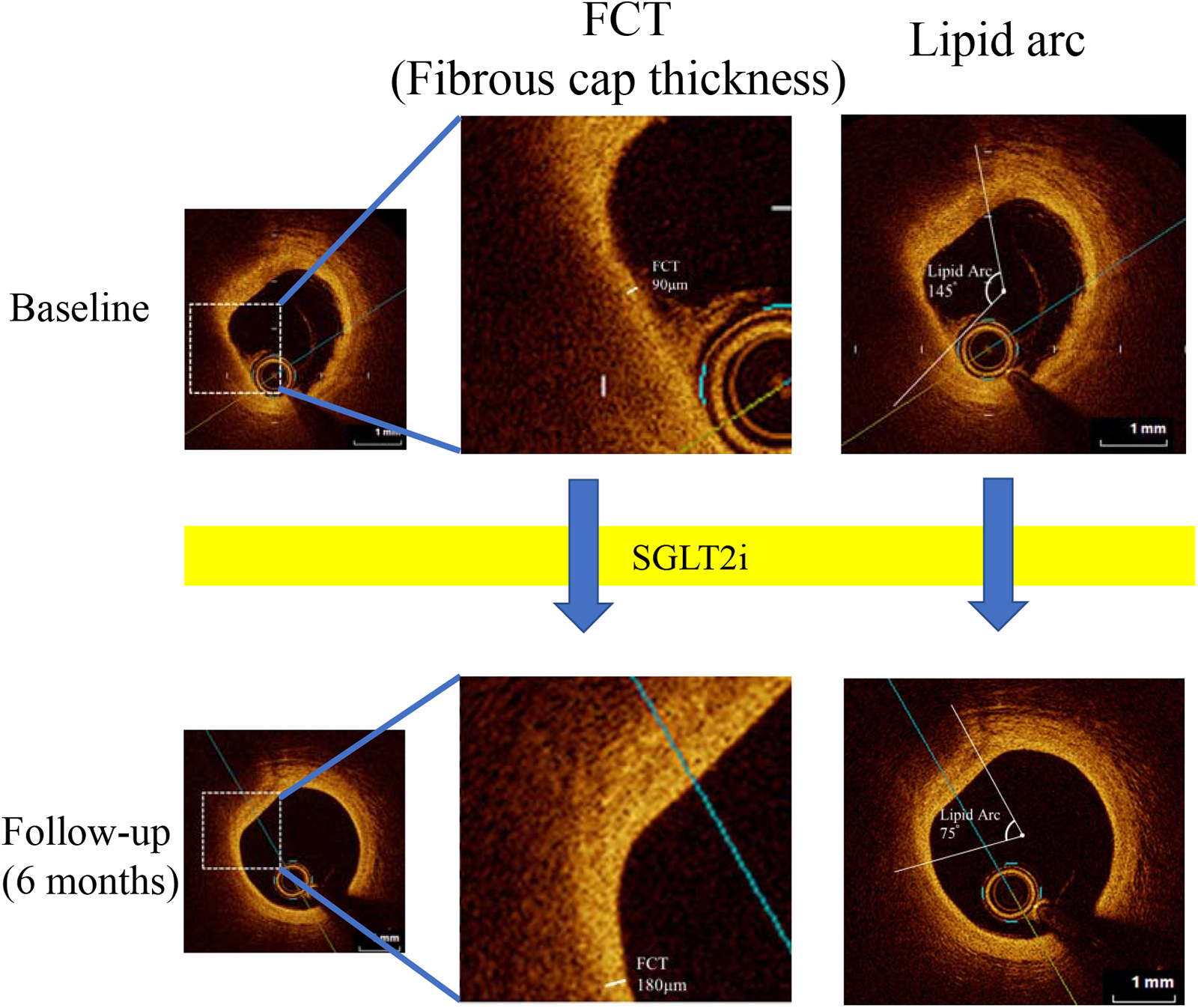
Representative images. The representative case showed NSTEMI and was performed PCI for LAD lesion. The OCT images had an unstable lesion at baseline (FCT 90 μm, Lipid arc 145°). The patient started to take SGLT2i from the hospitalization. After 6 months, OCT follow-up showed the lesion was improved (FCT 180 μm, Lipid arc 75°).

**Table 2.**
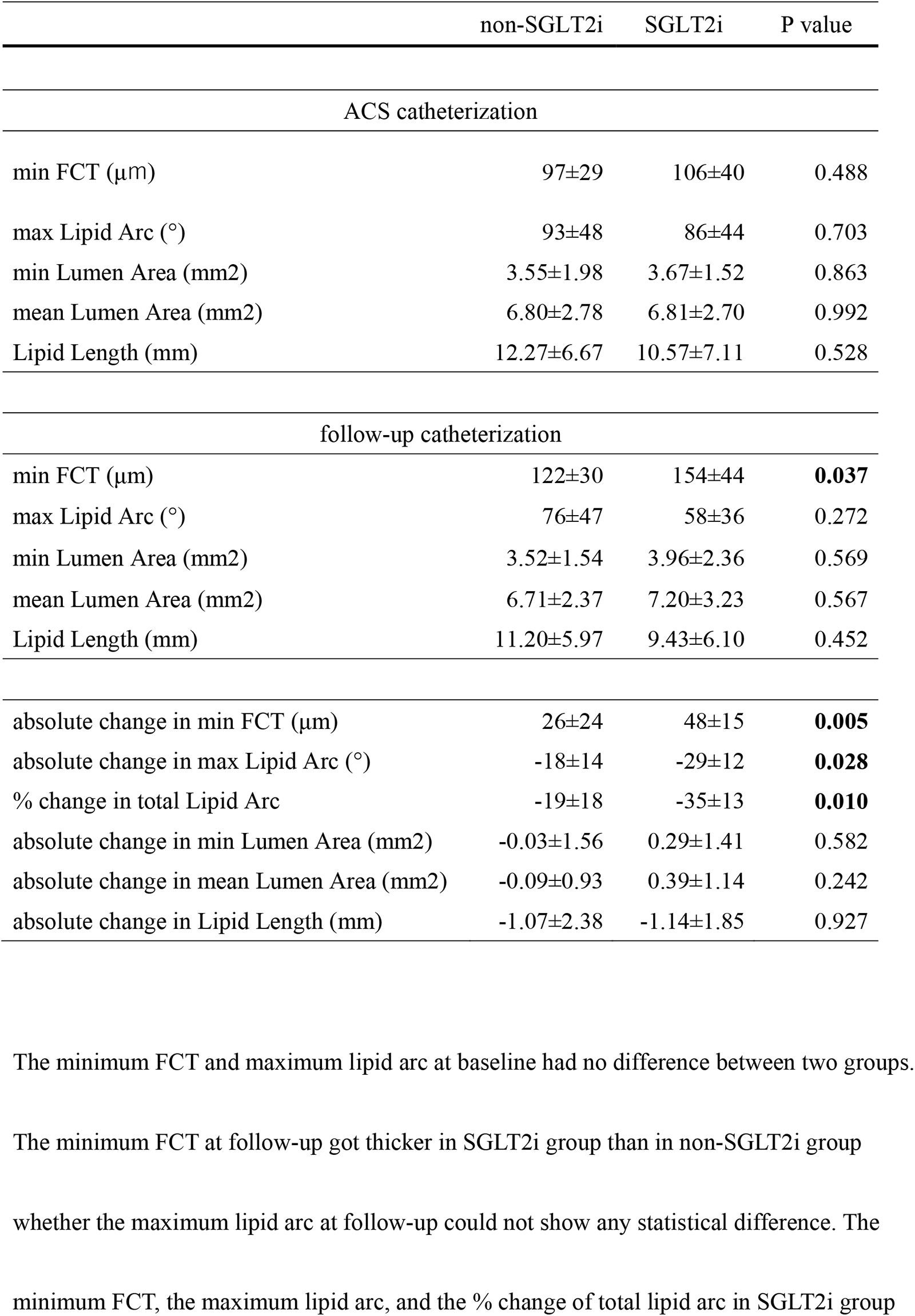
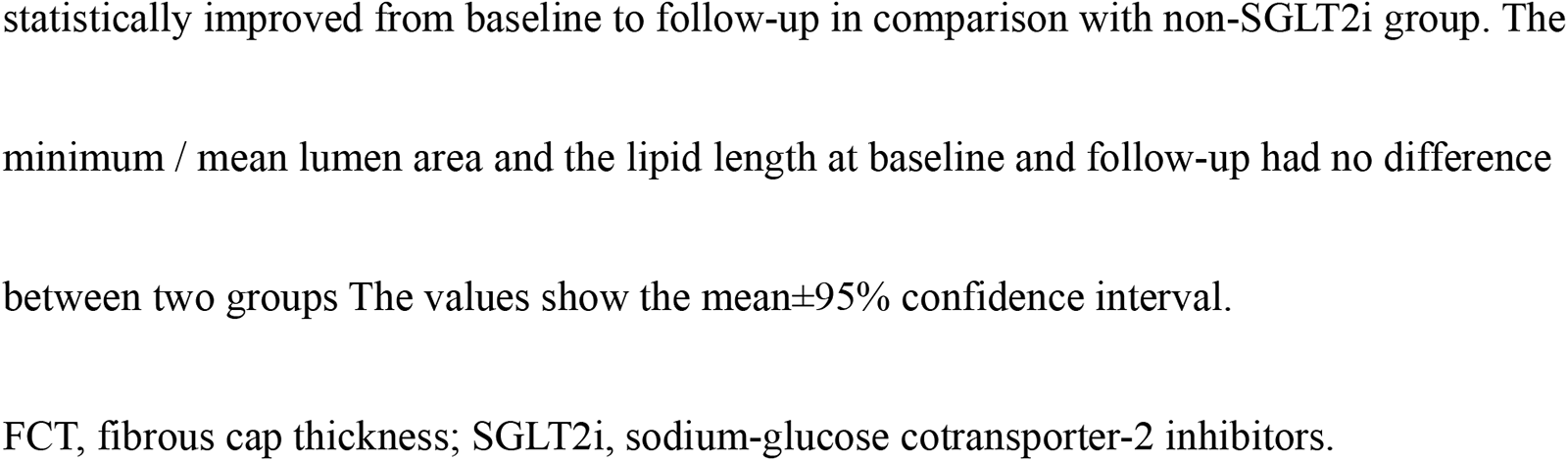
SGLT2i effect for unstable plaque.

The measurements of unstable plaque between the two cardiologists were comparable (Supplemental Table 1).

Table 3 shows the univariate analysis for FCT and lipid arc based on atherosclerosis-related variables, including sex, age, cardiovascular risks, medications, and laboratory data. None of these factors strongly correlated with minimum FCT or maximum lipid arc. SGLT2i use significantly correlated with the absolute change in the min FCT and max lipid arc.

**Table 3.**
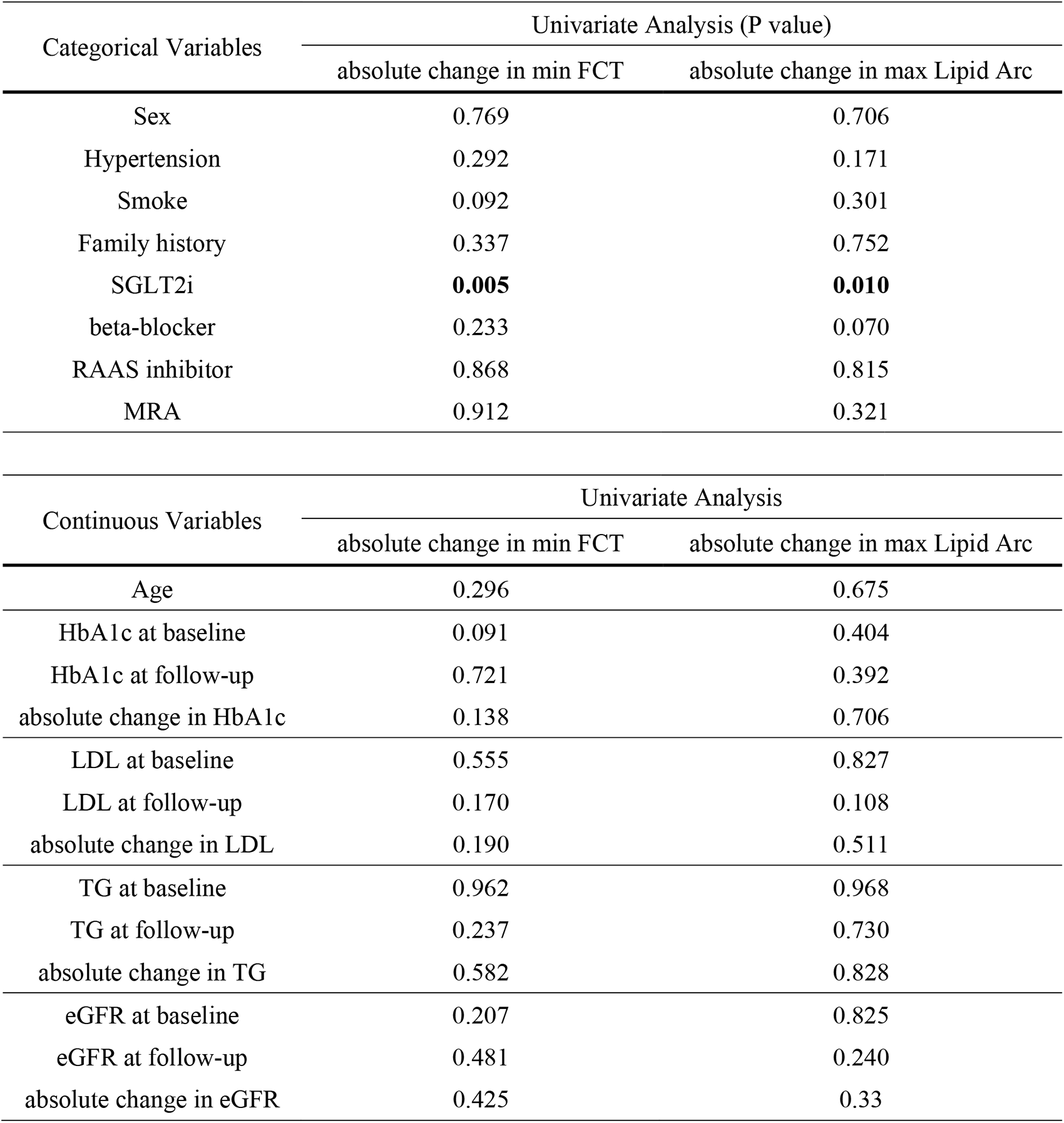
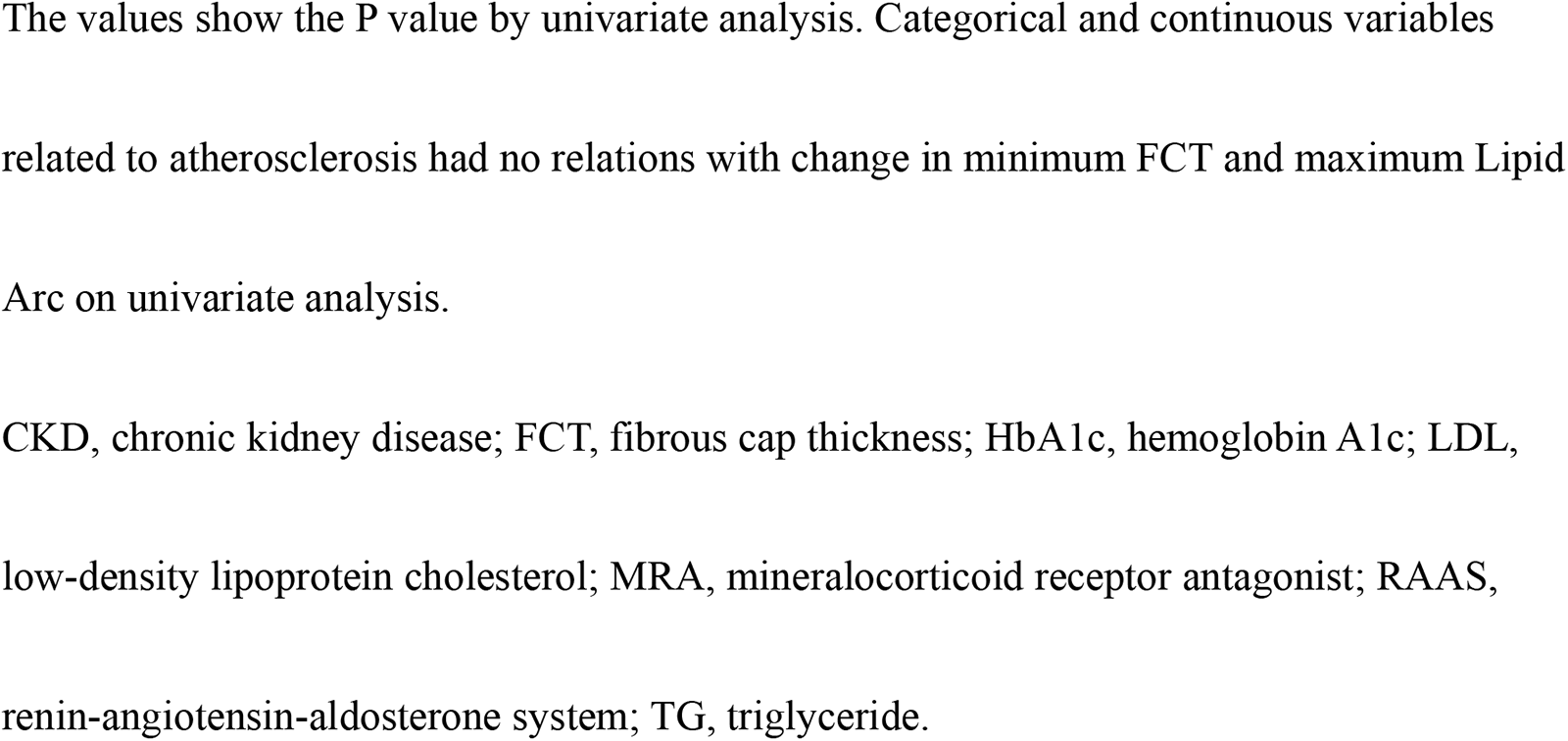
Univariate analysis with atherosclerosis-related variables.

### Clinical outcomes

To determine the effect of SGLT2i on clinical outcomes after ACS, we compared the incidence of MACE between the two groups for one year. Kaplan–Meier analysis showed a significantly lower incidence of MACE in the SGLT2i group than in the non-SGLT2i group (log-rank p = 0.023, unadjusted HR 4.72 [1.08, 20.63]) (Figure 4). Multivariable analysis of factors associated with MACE were TG value at hospitalization, SGLT2i, MRA, RAA inhibitor, anticoagulant drug, NSEMI, stent number and LVEF at baseline (Supplemental Table 2). The SGLT2i group showed a lower rate of all-cause mortality, revascularization due to ACS or coronary lesion progression, cerebrovascular disease, and hospitalization for heart failure but not significantly different (Supplemental Table 3). The SGLT2i group had a significantly lower revascularization rate after adjusting for age and HbA1c level at baseline (adjusted HR 6.77 [1.08, 42.52]).

**Figure 4.**
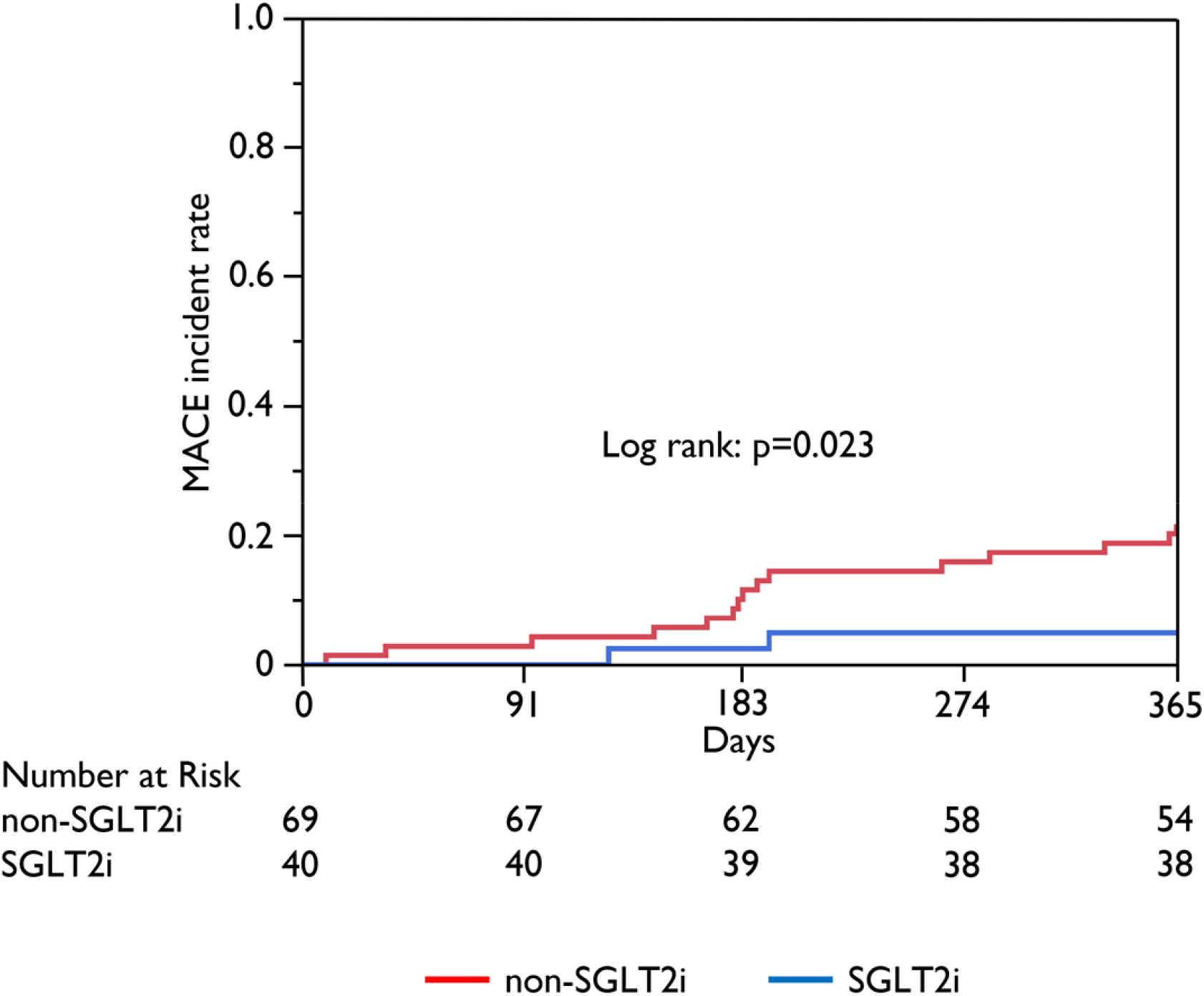
ACS prognosis and incidence rate of MACE. The cumulative incidence of MACE was evaluated using the Kaplan-Meier analysis. MACE included all-cause mortality, revascularization due to ACS or coronary lesion progression, cerebrovascular disease, and hospitalization for heart failure. The incidence of MACE was significantly lower in the SGLT2i group than in the non-SGLT2i group (log-rank test, p = 0.023).

## DISCUSSION

To the best of our knowledge, this is the first study to demonstrate the coronary lipid plaque-stabilizing effects of SGLT2i on ACS patients using coronary imaging. Regarding clinical events, SGLT2i reduced cardiovascular events after ACS. The main findings of this report are that: 1) in patients with T2DM after ACS, OCT analysis showed that the minimum FCT, the maximum lipid arc, and the % change of total lipid arc from baseline to follow-up improved significantly in the SGLT2i group, and 2) the patients with SGLT2i were associated with a lower clinical event rate, in particular lower subsequent coronary revascularizations, compared to those with non-SGLT2i.

### Effect of SGLT2i on cardiovascular prognosis

SGLT2i has been developed as an anti-diabetic drug that acts on Na-Glu channels to excrete sugar into the urine. SGLT2 inhibitors are one of four widely used drugs for heart failure, and several large randomized studies have reported that they reduce the risk of death from cardiovascular events or hospitalization for heart failure not only in patients with reduced ejection fraction but also in those with preserved ejection fraction.^9–12^ In addition, SGLT2i has been reported to be beneficial in patients with post-acute myocardial infarction, by improving cardiac function and reducing hospitalization for heart failure.^16–18^ Consistent with those previous findings, ^16–18^ we found that SGLT2i reduced clinical events and improved the prognosis of patients with ACS and T2DM after a one-year follow up.

### Coronary atherosclerosis in T2DM

Cholesterol-lowering agents, such as statin and ezetimibe, are the most common treatment to stabilize coronary atherosclerosis.^24, 25^ Moreover, additional PCSK9 inhibitor showed coronary plaque regression in patients of acute myocardial infarct even with statin.^26, 27^ However, even with these treatments, some coronary lesions may progress. Patients with T2DM are at high risk of coronary lesion progression.^1–3^ T2DM has been shown to accelerate the development of atherosclerosis and increase plaque instability/vulnerability, leading to ACS. The cholesterol-lowering management was reported not to be enough to control lipid plaque in patients with T2DM.^28^ In addition to the effects of SGLT2i on heart failure,^9–12^ SGLT2i not only has a primary preventive effect on AMI ^14, 15^ but also has secondary preventive effects on cardiovascular events after AMI.^16–18^ It was known that SGLT2 was overexpressed in the inflammatory process on atherosclerotic plaque of DM patients.^29^ SGLT2i can reduce or stabilize plaques by controlling the inflammatory cascade.^30–33^ Moreover, SGLT2i has been shown to improve insulin sensitivity in patients with T2DM,^34^ which could also have a cardiovascular protective effect.^35^ Their effects on atherosclerosis have been demonstrated in animal models ^19–21^ and in patients with stable ischemic heart disease ^22^, but not in patients with ACS.

### Effect of SGLT2i on coronary unstable plaque

Unstable plaques defined on OCT imaging, are known to increase the incidence of cardiovascular events such as ACS. The thickness of the fibrous cap, the size of the necrotic core, and the presence of macrophages indicate plaque instability.^36–38^ In this study, patients treated with SGLT2i had thicker fibrous caps and reduced lipid plaque arcs than those who did not receive SGLT2i. HbA1c value was worse in the SGLT2i group during hospitalization for ACS whether it was comparable at follow-up. The decrease of HbA1c value might be one of the reasons for plaque stabilization. However, the HbA1c level showed no correlation with the stabilization of unstable plaques in our results. As in previous studies,^22^ SGLT2i could stabilize unstable coronary plaque lesions in patients with ACS and T2DM, not by controlling diabetes, but by other mechanisms such as controlling inflammation in the hyperinflammatory cascade of ACS. Further investigation is required to elucidate the mechanism of action. Moreover, the SGLT2i group had lower LDL cholesterol, which could be one factor to improve unstable coronary lesions. However, it has been reported that diabetic patients are less likely to get plaque regression even with tight LDL control.^28^ No relationship between LDL values and unstable plaque was found in univariate analysis, suggesting that the effect of lower LDL is less than the effect of SGLT2i administration to stabilize coronary plaques. SGLT2i can affect unstable plaques even under lipid control.

We also found a reduction in the revascularization rate at one year after ACS. These findings suggest that histological improvements lead to better clinical outcomes. Therefore, SGLT2i administration should be considered in patients with ACS and T2DM undergoing PCI to stabilize coronary atherosclerotic lesions, in addition to improving cardiac function and controlling heart failure.

## STUDY LIMITATIONS

This study has some limitations. Firstly, it was a single-center, retrospective study. Secondly, the OCT cohort comprised a small patient group. Thirdly, core laboratories did not perform the OCT analysis. Finally, our physicians decided by themselves without consensus whether to administer SGLT2i to patients or not. Therefore, some of the baseline characteristics differed significantly between the SGLT2i group and non-SGLTi group. These differences might have influenced our study results. This is a pilot study to investigate SGLT2i on effective for coronary plaque. Prospective large-scale studies, including patients without diabetes, are required to overcome these limitations and make further progress in this area.

## CONCLUSIONS

SGLT2i administration for six months after ACS stabilized coronary lipid plaques as visualized by OCT imaging. It also reduced cardiovascular events one year after ACS. Our results suggest that SGLT2i improves atherosclerosis, which could explain the better prognosis of ACS. SGLT2i should, therefore, be used aggressively in patients with ACS and T2DM.

## Data Availability

The datasets generated and/or analyzed during the current study are available from the corresponding author on reasonable request.

## Abbreviations

ACS: Acute coronary syndrome
eGFR: Estimated glomerular filtration rate
FCT: Fibrous cap thickness
HbA1c: Hemoglobin A1c
LDL: Low-density lipoprotein cholesterol
MACE: Major adverse cardiovascular events
OCT: Optical coherence tomography
SGLT2i: Sodium-glucose cotransporter-2 inhibitors
TG: Triglyceride
T2DM: Type 2 diabetes mellites

## Acknowledgements

None.

## Supplemental Material

Figure S1

Table S1-S3

## REFERENCES

1. Cosentino F, Grant PJ, Aboyans V, Bailey CJ, Ceriello A, Delgado V, Federici M, Filippatos G, Grobbbee DE, Hansen TB, et al. 2019 ESC Guidelines on diabetes, pre-diabetes, and cardiovascular diseases developed in collaboration with the EASD. Eur Heart J. 2020;41:255–323.

2. Wilson S, Mone P, Kansakar U, Jankauskas SS, Donkor K, Adebayo A, Varzideh F, Eacobacci M, Gambardella J, Lombardi A, et al. Diabetes and restenosis. Cardiovasc Diabetol. 2022;21:23.

3. Godoy LC, Rao V, Farkouh ME. Coronary revascularization of patients with diabetes mellitus in the setting of acute coronary syndromes. Circulation. 2019;140: 1233–1235.

4. Zhang J, Zhang Q, Zhao K, Bian YJ, Liu Y, Xue YT. Risk factors for in-stent restenosis after coronary stent implantation in patients with coronary artery disease: A retrospective observational study. Medicine (Baltimore). 2022;101:e31707

5. Verma A, Patel AB, Waikar SS. SGLT2 Inhibitor: Not a traditional diuretic for heart failure. Cell Metab. 2020;32:13–14.

6. Zelniker TA, Braunwald E. Mechanisms of cardiorenal effects of sodium-glucose cotransporter 2 inhibitors: JACC State-of-the-Art Review. J Am Coll Cardiol. 2020;75:422–434.

7. Zinman B, Wanner C, Lachin JM, Fitchett D, Bluhmki E, Hantel S, Mattheus M, Devins T, Johansen OE, Woerle HJ, et al. Empagliflozin, cardiovascular outcomes, and mortality in Type 2 diabetes. N Engl J Med. 2015;373:2117–2128.

8. Neal B, Perkovic V, Mahaffey KW, Zeeuw DD, Fulcher G, Erondu N, Shaw W, Law G, Desai M, Matthews DR, et al. Canagliflozin and cardiovascular and renal events in Type 2 diabetes. N Engl J Med. 2017;377:644–657.

9. McMurray JJV, Solomon SD, Inzucchi SE, Køber L, Kosiborod MN, Martinez FA, Ponikowski P, Sabatine MS, Anand IS, Bělohlávek J, et al. Dapagliflozin in patients with heart failure and reduced ejection fraction. N Engl J Med. 2019;381:1995–2008.

10. Packer M, Anker SD, Butler J, Filippatos G, Ferreira JP, Pocock SJ, Sattar N, Brueckmann M, Jamal W, Cotton D, et al. Empagliflozin in patients with heart failure, reduced ejection Fraction, and volume overload: EMPEROR-reduced trial. J Am Coll Cardiol. 2021;77:1381–1392.

11. Anker SD, Butler J, Filippatos G, Ferreira JP, Bocchi E, Böhm M, Rocca HB, Choi D, Chopra V, Chuquiure-Valenzuela E, et al. Empagliflozin in heart failure with a preserved ejection fraction. N Engl J Med. 2021;385:1451–1461.

12. Bauersachs J. Heart failure drug treatment: the fantastic four. Eur Heart J. 2021;42:681–683.

13. Gallwitz B. The cardiovascular benefits associated with the use of sodium-glucose cotransporter 2 inhibitors – real-world Data. Eur Endocrinol. 2018;14:17–23.

14. Zhu J, Yu X, Zheng Y, Li J, Wang Y, Lin Y, He Z, Zhao W, Chen C, Qiu K, et al. Association of glucose-lowering medications with cardiovascular outcomes: an umbrella review and evidence map. Lancet Diabetes Endocrinol. 2020;8:192–205.

15. Zheng SL, Roddick AJ, Aghar-Jaffar R, Shun-Shin MJ, Francis D, Oliver N, Meeran K. Association between use of sodium-glucose cotransporter 2 inhibitors, glucagon-like peptide 1 agonists, and dipeptidyl peptidase 4 inhibitors with all-cause mortality in patients with Type 2 diabetes: A systematic review and meta-analysis. JAMA. 2018;319:1580–1591.

16. Zhu Y, Zhang JL, Yan XJ, Sun L, Ji Y, Wang FF. Effect of dapagliflozin on the prognosis of patients with acute myocardial infarction undergoing percutaneous coronary intervention. Cardiovasc Diabetol. 2022;21:186.

17. Harrington J, Udell JA, Jones WS, Anker SD, Bhatt DL, Petrie MC, Vedin O, Sumin M, Zwiener I, Hernandez AF, et al. Empagliflozin in patients post myocardial infarction rationale and design of the EMPACT-MI trial. Am Heart J. 2022;253:86–98.

18. Lewinski DV, Kolesnik E, Tripolt NJ, Pferschy PN, Benedikt M, Wallner M, Alber H, Berger R, Lichtenauer M, Saely CH, et al. Empagliflozin in acute myocardial infarction: the EMMY trial. Eur Heart J. 2022;43:4421–4432.

19. Terasaki M, Hiromura M, Mori Y, Kohashi K, Nagashima M, Kushima H, Watanabe T, Hirano T. Amelioration of hyperglycemia with a sodium-glucose cotransporter 2 inhibitor prevents macrophage-driven atherosclerosis through macrophage foam cell formation suppression in type 1 and type 2 diabetic mice. PLoS One. 2015;10: e0143396.

20. Liu Y, Xu J, Wu M, Xu B, Kang L. Empagliflozin protects against atherosclerosis progression by modulating lipid profiles and sympathetic activity. Lipids Health Dis. 2021;20:5.

21. Chen Y, Jandeleit-Dahm K, Peter K. Sodium-Glucose Co-Transporter 2 (SGLT2) Inhibitor Dapagliflozin Stabilizes Diabetes-Induced Atherosclerotic Plaque Instability. J Am Heart Assoc. 2022;11(1):e022761.

22. Sardu C, Trotta MC, Sasso FC, Sacra C, Carpinella G, Mauro C, Minicucci F, Calabrò P, Amico MD’, Ascenzo FD’, et al. SGLT2-inhibitors effects on the coronary fibrous cap thickness and MACEs in diabetic patients with inducible myocardial ischemia and multi vessels non-obstructive coronary artery stenosis. Cardiovasc Diabetol. 2023;22(1):80.

23. American Diabetes Association. 2. Classification and diagnosis of diabetes: Standards of medical care in diabetes-2021. Diabetes Care. 2021;44(Suppl.1):S15–33.

24. Komukai K, Kubo T, Kitabata H, Matsuo Y, Ozaki Y, Takarada S, Okumoto Y, Shiono Y, Orii M, Shimamura K, et al. Effect of Atorvastatin Therapy on Fibrous Cap Thickness in Coronary Atherosclerotic Plaque as Assessed by Optical Coherence Tomography: The EASY-FIT Study. J Am Coll Cardiol. 2014;64(21):2207–17.

25. Habara M, Nasu K, Terashima M, Ko E, Yokota D, Ito T, Kurita T, Teramoto T, Kimura M, Kinoshita Y, et al. Impact on optical coherence tomographic coronary findings of fluvastatin alone versus fluvastatin + ezetimibe. Am J Cardiol. 2014;15;113(4):580–7.

26. Räber L, Ueki Y, Otsuka T, Losdat S, Häner JD, Lonborg J, Fahrni G, Iglesias JF, van Geuns R, Ondracek AS, et al. Effect of alirocumab added to high-intensity statin therapy on coronary atherosclerosis in patients with acute myocardial infarction: the PACMAN-AMI randomized clinical trial. JAMA. 2022;327(18):1771–81.

27. Nicholls SJ, Kataoka Y, Nissen SE, Prati F, Windecker S, Puri R, Hucko T, Aradi D, Herrman JR, Hermanides RS, et al. Effect of evolocumab on coronary plaque phenotype and burden in statin-treated patients following myocardial infarction. JACC Cardiovasc Imaging. 2022;15(7):1308–21.

28. Iwai T, Kataoka Y, Nicholls SJ, Puri R, Murata S, Nishimura K, Murai K, Kitahara S, Sawada K, Matama H, et al. Phenotypic Features of Coronary Atheroma in Diabetic and Nondiabetic Patients With Low-Density Lipoprotein Cholesterol &<55 mg/dL. (letter) JACC Cardiovasc Imaging. 2022;15(6):1166–1169.

29. D’Onofrio N, Sardu C, Trotta MC, Scisciola L, Turriziani F, Ferraraccio F, Panarese I, Petrella L, Fanelli M, Modugno P, et al. Sodium–glucose co-transporter2 expression and inflammatory activity in diabetic atherosclerotic plaques: effects of sodium–glucose co-transporter2 inhibitor treatment. Mol Metab. 2021;54:101337.

30. Mortanges AP, Salvador D, Laimer M, Muka T, Wilhelm M, Bano A. The role of SGLT2 inhibitors in atherosclerosis: A narrative mini-review. Front Pharmacol. 2021;12:751214.

31. Cowie MR, Fisher M. SGLT2 inhibitors: mechanisms of cardiovascular benefit beyond glycaemic control. Nat Rev Cardiol. 2020;17:761–772.

32. Elrakaybi A, Laubner K, Zhou Q, Hug MJ, Seufert J. Cardiovascular protection by SGLT2 inhibitors–-Do anti-inflammatory mechanisms play a role? Mol Metab. 2022;64:101549.

33. Spigoni V, Fantuzzi F, Carubbi C, Pozzi G, Masselli E, Gobbi G, Solini A, Bonadonna RC, Cas AD. Sodium-glucose cotransporter 2 inhibitors antagonize lipotoxicity in human myeloid angiogenic cells and ADP-dependent activation in human platelets: potential relevance to prevention of cardiovascular events. Cardiovasc Diabetol. 2020;19:46.

34. Merovci A, Herrera CS, Daniele G, Eldor R, Fiorentino TV, Tripathy D, Xiong J, Perez Z, Norton L, Abdul-Ghani MA, et al. Dapagliflozin improves muscle insulin sensitivity but enhances endogenous glucose production. J Clin Invest. 2014; 124:509–514.

35. Hill MA, Yang Y, Zhang L, Sun Z, Jia G, Parrish AR, Sowers JR. Insulin resistance, cardiovascular stiffening and cardiovascular disease. Metabolism. 2021;119:154766.

36. Sinclair H, Bourantas C, Bagnall A, Mintz GS, Kunadian V. OCT for the identification of vulnerable plaque in acute coronary syndrome. JACC Cardiovasc Imaging. 2015;8:198–209.

37. Russo M, Kim HO, Kurihara O. Characteristics of non-culprit plaques in acute coronary syndrome patients with layered culprit plaque. Eur Heart J Cardiovasc Imaging. 2020;21:1421–1430.

38. van Veelen A, van der Sangen NMR, Henriques JPS, Claessen BEPM. Identification and treatment of the vulnerable coronary plaque. Rev Cardiovasc Med. 2022;23:39.

